# Transmission of rifampicin-resistant tuberculosis in Ho Chi Minh City, Viet Nam: a prospective genomic epidemiology study

**DOI:** 10.64898/2026.03.21.26348963

**Authors:** Ruan Spies, Nguyen Hong Hanh, Phan Trieu Phu, Luong Kim Lan, Kim Lan, Ngo Ngoc Hue, Nguyen Le Quang, Do Dang Anh Thu, Nguyen Thi Le Huong, Tran Le Thi Ngoc Thao, Trinh Thi Bich Tram, Vu Thi Ngoc Ha, Dang Thi Minh Ha, Nguyen Huu Lan, Nguyen Phuc Hai, Nguyen Hung Thuan, Tran Thi Kim Quy, Viola Dreyer, Stefan Niemann, Derrick Crook, Le Hong Van, Guy Thwaites, Nguyen Thuy Thuong Thuong, Marc Choisy, James Watson, Timothy Walker

**Affiliations:** Oxford University Clinical Research Unit, Ho Chi Minh City, Viet Nam; Centre for Tropical Medicine and Global Health, Nuffield Department of Medicine, University of Oxford, United Kingdom; Pham Ngoc Thach Hospital, Ho Chi Minh City, Viet Nam; German Center for Infection Research (DZIF), Partner Site Hamburg-Lübeck-Borstel-Riems, Borstel, Germany; Research Center Borstel, Leibniz Lung Center, Molecular and Experimental Mycobacteriology, Borstel, Germany; Research Center Borstel, Leibniz Lung Center, Supranational and National Reference Center for Mycobacteria, Borstel, Germany; Modernising Medical Microbiology Unit, Nuffield Department of Medicine, University of Oxford, United Kingdom; Shared Hospital Laboratory and Sunnybrook Research Institute, Sunnybrook Hospital, Toronto, Canada; Infectious Diseases Data Observatory, Oxford, United Kingdom

## Abstract

**Background:** Rifampicin-resistant tuberculosis (RR-TB) is a major threat to public health in Viet Nam, with nearly 10,000 incident cases estimated annually. It is uncertain whether these cases are driven by transmission of resistant strains or *de novo* resistance acquisition during treatment.

**Methods:** We undertook dense, city-wide sampling of adults newly diagnosed with pulmonary RR-TB in Ho Chi Minh City, Viet Nam’s largest city, between March 2020 and April 2024. Participants provided sputum for culture and whole-genome sequencing (WGS), and demographic and clinical data were collected at enrolment. Phylogenetic analyses were combined with clinical histories to infer transmitted versus acquired rifampicin resistance. Estimates were corrected for sampling coverage using simulation-extrapolation (SIMEX). Temporal emergence of rifampicin resistance was reconstructed by lineage using Bayesian phylogenetic dating, and the geographic and demographic structure of transmission networks was assessed using geocoded residential data and commute time-based analyses.

**Findings:** Among 2,319 RR-TB cases diagnosed during the study period, 1,491 (64%) isolates were successfully sequenced. After accounting for sampling and phylogenetic uncertainty, we estimated that between 72% and 87% of all RR-TB arose through transmission of already-resistant strains with the remainder due to *de novo* acquired resistance. Bayesian dating analyses revealed that resistance emergence events occurred repeatedly from the 1980s to the present, with early events seeding long-lived, city-wide transmission networks. Transmission networks were geographically dispersed across the city, with limited household clustering, and only weakly structured by host demographics, consistent with diffuse, city-wide transmission rather than localised or assortative spread.

**Interpretation:** RR-TB in Ho Chi Minh City is driven predominantly by ongoing transmission, but a substantial minority of cases arise from newly acquired resistance. Alongside promoting early diagnosis and treatment to interrupt transmission, the main drivers of acquired resistance need to be identified to control RR-TB.

**Funding:** The Rhodes Trust, Wellcome.

**Research in context:** *Evidence before this study:* We searched PubMed from database inception up to November 2025 for studies describing the transmission and acquisition of multidrug /rifampicin-resistant tuberculosis (MDR/RR-TB), using terms including “tuberculosis”, “rifampicin resistance”, “multidrug resistance”, “genomic epidemiology”, “whole-genome sequencing”, “transmission” and “acquired resistance”. We additionally added terms “Ho Chi Minh City” and “Viet Nam” to identify results specific our study setting. Previous studies from high burden settings typically reported that MDR/RR-TB isolates clustered with high proportions, suggesting transmission is the major contributor to MDR/RR-TB burden. However, estimates varied widely across settings, study designs and analytic approaches. Many studies relied on SNP-based clustering thresholds and did not account for incomplete sampling. Few studies integrated treatment history and homoplasy-aware phylogenetic inference or accounted for uncertainty in their estimates of the population-level contribution of *de novo* rifampicin resistance acquisition. No studies from Viet Nam addressed this question and the extent to which transmitted and acquired resistance contribute to the burden of MDR/RR-TB remains unknown.

*Added value of this study:* We combined dense, city-wide sampling of RR-TB in a high-burden metropolitan setting with whole-genome sequencing, historic genomic context and phylogenetic analyses to quantify the contributions of transmission and *de novo* acquisition to MDR/RR-TB burden. By integrating phylogenetic placement of resistance mutations with individual treatment history, we provide robust population-level estimates of transmission and independent resistance emergence while explicitly accounting for phylogenetic and sampling uncertainty. Acknowledging the potential effect of sampling coverage on these estimates, we further provide estimates extrapolated to complete coverage using a simulation-extrapolation framework. We show that the majority of RR-TB in Ho Chi Minh City is attributable to transmission, but that a substantial minority of cases reflect *de novo* acquisition of rifampicin resistance. Our findings demonstrate that *de novo* resistance acquisition continues to occur notwithstanding improvements in programmatic TB control, treatment success rates, and diagnostic capacity.

*Implications of all of the available evidence:* Our findings emphasise that MDR/RR-TB in Ho Chi Minh City is overwhelmingly driven by transmission and that controlling it requires a transmission-focused response. Intensified efforts to interrupt transmission through earlier case detection and rapid treatment initiation should be programmatic priorities. Concurrently, further investigation into the precise drivers of acquired drug resistance, which represents a potentially preventable fraction of the RR-TB burden, remains warranted

## Introduction

The global incidence of tuberculosis (TB), including rifampicin-resistant TB (RR-TB), is falling. However, the pace of decline remains insufficient to meet the World Health Organization’s End TB targets.^1^ In Viet Nam, TB incidence mirrors the global downward trend, but the burden of RR-TB has remained largely unchanged over the last decade, with nearly 10,000 new cases estimated annually.^1^ Historical data from Ho Chi Minh City, the country’s largest city, indicate that most people diagnosed with RR-TB have had a previous episode of TB.^2^ This may be because RR-TB is only more recently being correctly diagnosed, or because resistance has been selected through inadequate treatment. Either way, the scale of the public health problem underscores the importance of understanding what is driving RR-TB in this setting.

Central to this question is distinguishing between the two pathways by which rifampicin resistance can arise: transmission of resistant strains or *de novo* acquisition of resistance mutations during treatment for rifampicin-susceptible disease. This distinction has direct public health relevance, as each pathway requires different interventions. Interrupting transmission requires interventions that facilitate rapid diagnosis and treatment, such as active case finding,^3^ whereas preventing acquired resistance depends on patients receiving the right number of the right drugs, at the right dose, for the right duration.^4^

In the early years of the drug-resistant TB epidemic, rifampicin resistance was believed to arise mainly from inadequate treatment, and resistant strains were thought to carry fitness costs that limited their capacity for widespread transmission.^5^ The DOTS strategy, standardised treatment regimens, and fixed-dose drug combinations were designed to prevent such acquired resistance.^6^ As these measures became established, our understanding of the epidemic evolved, with molecular epidemiology and modelling studies subsequently demonstrating that resistant *Mycobacterium tuberculosis* strains transmit effectively, and that transmission of already-resistant strains likely accounts for most RR-TB globally.^7,8^

Viet Nam’s mature National TB Programme (NTP) includes long-standing safeguards against acquired resistance including DOTS, fixed-dose drug combinations and uninterrupted access to 1^st^ line drugs^9^, suggesting the contemporary RR-TB burden should be driven predominantly by transmission of established resistant clones. We conducted comprehensive sampling and whole-genome sequencing (WGS) of rifampicin-resistant *M. tuberculosis* isolates across Ho Chi Minh City over a four-year period to test this hypothesis and inform targeted control strategies by identifying risk-factors for transmission.

## Methods

### Study setting and design

Ho Chi Minh City has a population of approximately 10 million people and a high burden of TB and RR-TB.^10^ All individuals diagnosed with RR-TB in Ho Chi Minh City are referred to initiate treatment at Phạm Ngọc Thạch Hospital, the city’s specialist centre for TB and lung disease, which provides treatment for approximately 80% of drug-resistant TB cases in Viet Nam.^11^

We conducted a prospective cohort study aiming to enrol all adult residents (≥18 years) newly diagnosed with pulmonary RR-TB by Xpert MTB/RIF between March 2020 and May 2024. Eligible participants provided written informed consent at their first appointment at Phạm Ngọc Thạch Hospital. At enrolment, and prior to initiating treatment, participants submitted a sputum specimen for culture and WGS and completed a standardised case-report form recording demographic characteristics and clinical history.

This study received ethical approval from the Institutional Review Board at Phạm Ngọc Thạch Hospital (643/PNT-HDDD) and the Oxford Tropical Research Ethics Committee (reference no. 51–19).

### Sample processing

Sputum sample processing and culture have been described previously.^12^ Genomic DNA was extracted from cultured isolates using the cetyl trimethylammonium bromide (CTAB) method^13,14^ and prepared for sequencing using a modified Nextera XT library preparation protocol^15^, generating paired-end reads on Illumina NextSeq 1000/2000 platforms with a target mean depth of ≥50X **(Supplementary Methods)**. Bioinformatic processing was performed using the GPAS *Mycobacterium* pipeline v2.2.1 to determine species and lineage, predict drug resistance and generate intermediate files for phylogenetic analyses^16^.

### Phylogenetic analysis

To place the newly sequenced isolates in evolutionary context, we combined them with previously published *M.*L*tuberculosis* genomes from Ho Chi Minh City and additional genomes we generated in a linked contemporaneous study **(Supplementary Methods)**. This combined dataset included both rifampicin-resistant and rifampicin-susceptible isolates, providing the context required to identify the emergence of resistance mutations. Maximum-likelihood phylogenies were inferred using IQ-TREE v3.0.1 **(Supplementary Methods)**. Samples with evidence of mixed-lineage infection, defined by the detection of lineage-specific SNPs from more than one *M. tuberculosis* sub-lineage, were excluded because their phylogenetic placement could not be determined with confidence.

We classified rifampicin resistance as arising through transmission (primary resistance) or *de novo* acquisition (secondary resistance) by first looking at the clinical history and then the phylogenetic placement of resistance-conferring mutations. Participants with no prior TB were classified as having developed RR-TB through transmission, regardless of phylogenetic position, as they had no opportunity for treatment-driven resistance selection. Among participants with previous TB, classification was determined by how resistance mutations mapped to branches in the phylogeny **(Figure S1)**. To this end, we identified and mapped mutations in the *rpoB* gene that had arisen independently across multiple branches of the phylogeny using SNPPar^21^ and annotated these according to the World Health Organisation mutation catalogue.^22^ Where a resistance mutation mapped to the branch leading to a single isolate, we assumed the most parsimonious interpretation that resistance emerged in that individual *de novo* rather than being inherited through transmission from an ancestor.

When a resistance mutation mapped to an internal node in the phylogeny there was no means of determining whether all sampled descendant strains inherited it through transmission or whether it actually emerged *de novo* in one of those strains with subsequent transmission to the other decedents. We therefore undertook two alternative analyses: the first (hereafter ‘N’) classified all N descendants of an internal resistance node as having developed RR-TB through transmission, providing a lower bound on the number of *de novo* acquired resistance events. The second approach (hereafter ‘N −1’) attributed *de novo* acquired resistance to one descendant strain so long as at least one of the patients from whom a descendant strain was obtained had a history of prior TB. This approach makes the parsimonious assumption that the remaining N −1 descendants inherited resistance through transmission **(Figure S1)**. Together, these approaches defined plausible ranges for the proportion of RR-TB arising through transmission and *de novo* acquisition.

### Uncertainty and sensitivity analyses

We implemented a bootstrap procedure to account for uncertainty from both sampling variability and phylogenetic inference. In each of 100 iterations, the full dataset was resampled with replacement. Because phylogenetic inference requires unique sequences, trees were constructed using the unique set of resampled isolates, while the original resampling weights were retained for estimation.

Classification of transmitted versus acquired resistance was performed under both the N and N-1 approaches, with each sample’s contribution weighted by its resampling frequency. Convergence was assessed at both population and sample levels by monitoring the running mean of the proportion of acquired resistance across replicates and the stability of individual sample classification probabilities. The proportion of acquired RR-TB was summarised as the plausible range spanned by the two classification approaches, with empirical 95% confidence intervals derived from the 2.5th and 97.5th percentiles of the bootstrap distributions.

Because apparent independent emergence could actually be the consequence of an infection with a resistant strain from outside Ho Chi Minh City (importation) rather than true *de novo* acquisition, we conducted a sensitivity analysis with respect to birthplace. Among participants with a history of previous TB treatment, we used fractional logistic regression to estimate the odds of acquired (versus transmitted) rifampicin resistance by birthplace under the N approach definition from the bootstrap-derived probability of acquired resistance. Using the same approach, we also conducted exploratory univariate analyses of potential risk factors for acquired resistance, including *M. tuberculosis* lineage, age, sex, HIV status, diabetes, BMI, smoking, alcohol use, and intravenous drug use.

### Correction for incomplete sampling

Incomplete sampling may bias classification towards a label of *de novo* acquired resistance. When cases are missing from the dataset, true transmission networks may be fragmented, causing mutations that arose on shared internal branches to instead appear as independent events. To address this, we applied a simulation-extrapolation (SIMEX) framework.^23^ We simulated progressively sparser sampling of the RR-TB isolates collected in this study (for which we were able to estimate sampling coverage) by subsampling the dataset at ten evenly-spaced fractions, retaining between 8% and 92% of the sampled isolates. We rebuilt phylogenies at each fraction and used generalised additive models (GAMs) to characterise how the estimated proportion of *de novo* acquired resistance changed as fewer cases were sampled. We then used the fitted GAMs to extrapolate to 100% sampling coverage. To further assess whether estimates were also sensitive to the temporal depth of sampling (i.e., if estimates would differ had sampling begun earlier than 2020), we repeated the analysis holding sample size constant while varying the range of years sample from one to five **(Supplementary Methods)**.

### Temporal emergence of rifampicin resistance

To estimate when rifampicin resistance emerged in the major *M.*L*tuberculosis* lineages circulating in Viet Nam (Lineages 1, 2, and 4), we performed Bayesian phylogenetic dating using BactDating.^24^ We analysed each lineage separately to allow for lineage-specific mutation rates. For Lineages 1 and 2, molecular clock rates were estimated from the data using a subsampling approach; for Lineage 4, we applied a previously published rate (0.237 substitutions/genome/year)^25^ as there was insufficient temporal signal to reliably estimate a context-specific rate **(Supplementary Methods)**. The timing of resistance emergence was inferred from the estimated age of the most recent common ancestor of each resistant clade.

### Geographic and demographic structure of transmission networks

Transmission networks were defined as groups of isolates descending from a shared internal node where a rifampicin-resistance mutation had arisen. Residential addresses for participants from the present study were geocoded using the Google Maps Geocoding API via the *tidygeocoder* R package.^26^ To assess whether household transmission was a major driver of RR-TB, we identified cases sharing the same residential address and examined whether co-resident cases belonged to the same or different transmission networks.

We characterised the structure of RR-TB transmission along three dimensions. To assess whether transmission was geographically localised or dispersed, we mapped transmission networks, measured the dispersion of each network using median pairwise commute times between cases derived from the Open-Source Routing Machine (OSRM)^27^, and tested whether genetically related cases lived closer together than unrelated cases using the Mantel test. Dispersion was quantified for networks with ≥2 geocoded cases, and the association between dispersion and estimated year of emergence was assessed using Spearman’s rank correlation.

Within- versus between-network commute times were compared using the Wilcoxon rank-sum test. To assess whether transmission was occurring preferentially between individuals sharing demographic characteristics (assortative transmission), we used treeSeg^28^, a multiscale change-point approach that tests for dependence between a trait and the phylogenetic tree structure at all levels of the tree hierarchy simultaneously while controlling the false positive rate at a prespecified level (α = 0.05). Binary traits (sex, HIV status, diabetes, birthplace [Ho Chi Minh City vs elsewhere] and drug use history) were tested directly and age was tested as a continuous variable under a Gaussian model. Demographic distributions were also compared across large transmission networks (≥10 members). To quantify the joint explanatory value of geographic proximity and demographic similarity for genetic relatedness, we fitted multiple regression on distance matrices (MRM) **(Supplementary Methods)**.

## Results

Between March 1^st^, 2020 and April 30^th^, 2024, 2,596 Ho Chi Minh City residents were diagnosed with pulmonary RR-TB by routine services using Xpert MTB/RIF. We enrolled 2,033 (78%) of these individuals into our prospective cohort study. Reasons for non-enrolment were not formally recorded but were largely attributed to temporary recruitment interruptions during COVID-19 lockdowns. Among enrolled participants, 1,698 (84%) had sputum samples successfully cultured and whole-genome sequenced. Among these, WGS data indicated that 108 (6%) were rifampicin-susceptible and that 74 (4%) were non-tuberculous mycobacteria (NTM). 25 (1%) had insufficient sequence coverage for resistance prediction **(Figure 1)**. We considered the rifampicin-susceptible and NTM isolates (182/1,698) to represent programmatic false positives. Applying this 10.7% false positive rate to the entire RR-TB population, we estimated that we successfully sequenced 1,491/2,319 (64%) of all true RR-TB cases during the study period. Participants without a sequenced isolate did not differ from those with sequencing data, apart from a higher proportion with smear-negative disease **(Table S1)**.

**Figure 1:**
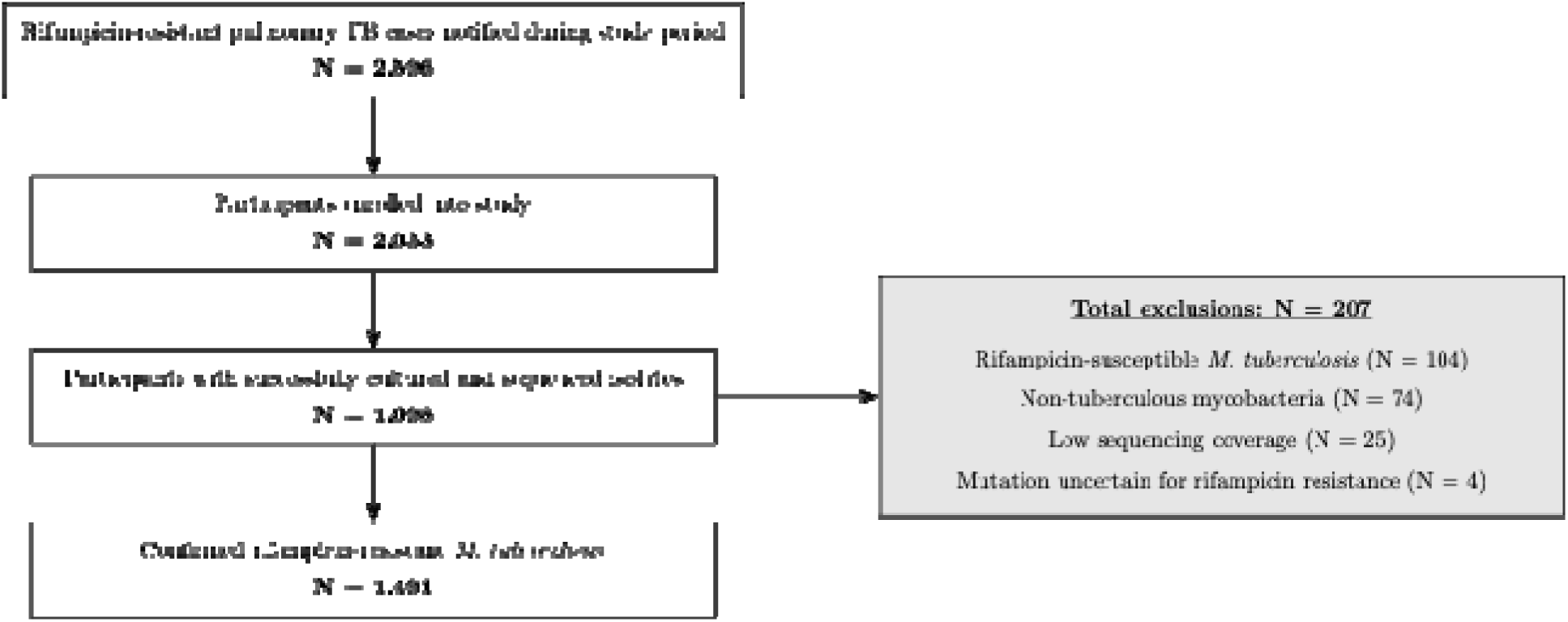
**Study flow diagram** 1,070 (72%) participants were male, mean age was 46 and mean BMI 19.5. 802 (54%) participants had previous TB and 435 (29%) self-reported a diagnosis of diabetes. Participants lived in households with an average of 3.6 additional members **(Table 1)**. Most isolates were Lineage 2 [1,320 (89%)] (Figure 2).

**Figure 2:**
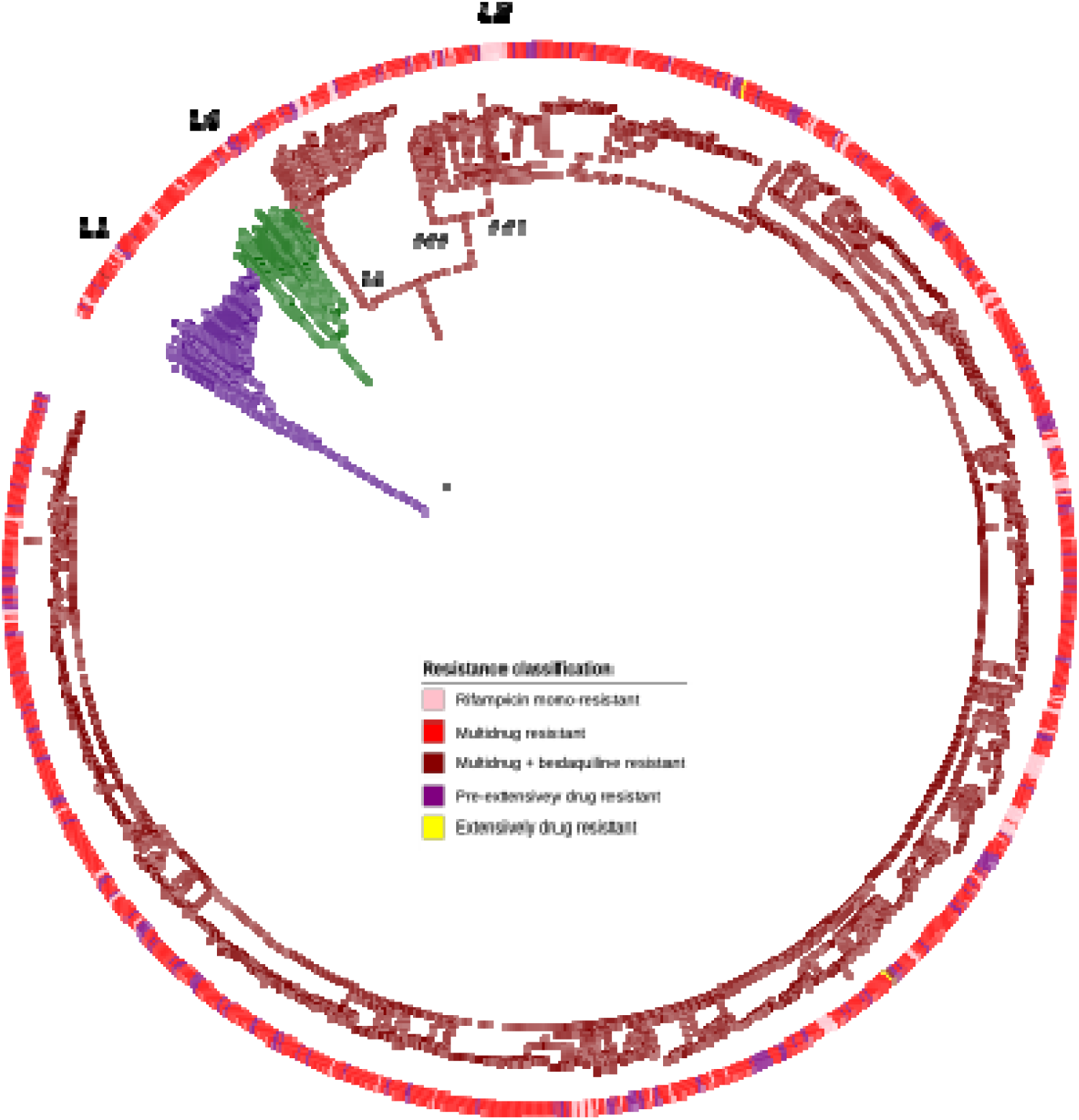
**Maximum-likelihood phylogeny for rifampicin-resistant M. tuberculosis isolates** *\*Excludes mixed lineage isolates. Outer ring represents drug resistance classification and branch colours represent lineage*

**Table 1:** Participant characteristics stratified by TB history.

|  | Overall<br>(N=1491) | Previous TB<br>(N=802) | No Previous TB<br>(N=689) |
| --- | --- | --- | --- |
| **Age, mean (SD)** | 46 (14) | 48 (14) | 43 (15) |
| **Male sex, n** | 1070 (71.8%) | 625 (77.9%) | 445 (64.6%) |
| **Body mass index, mean (SD)** | 19.5 (3.20) | 19.2 (3.18) | 19.7 (3.20) |
| **Additional people in household, mean (SD)** | 3.60 (2.56) | 3.62 (2.66) | 3.59 (2.45) |
| **HIV positive, n** | 90 (6.0%) | 67 (8.4%) | 23 (3.3%) |
| **Diabetes, n** | 435 (29.2%) | 249 (31.0%) | 186 (27.0%) |
| **Smoking status, n** |  |  |  |
| Never | 669 (44.9%) | 295 (36.8%) | 374 (54.3%) |
| Previous | 499 (33.5%) | 311 (38.8%) | 188 (27.3%) |
| Current | 322 (21.6%) | 195 (24.3%) | 127 (18.4%) |
| **Alcohol consumption, n** |  |  |  |

|  | **Overall**<br>**(N=1491)** | **Previous TB**<br>**(N=802)** | **No Previous TB**<br>**(N=689)** |
| --- | --- | --- | --- |
| Never | 752 (50.4%) | 375 (46.8%) | 377 (54.7%) |
| <14 units/week | 424 (28.4%) | 235 (29.3%) | 189 (27.4%) |
| >14 units/week | 315 (21.1%) | 192 (23.9%) | 123 (17.9%) |
| **Previous intravenous drug use, n** | 47 (3.2%) | 39 (4.9%) | 8 (1.2%) |
| **Smear microscopy positive, n** | 1138 (76.3%) | 625 (77.9%) | 513 (74.5%) |
| **Resistance classification, n** |  |  |  |
| Rifampicin mono-resistant | 154 (10.3%) | 80 (10.0%) | 74 (10.7%) |
| Multidrug resistant | 1110 (74.4%) | 585 (72.9%) | 525 (76.2%) |
| Pre-extensively drug resistant | 221 (14.8%) | 132 (16.5%) | 89 (12.9%) |
| Multidrug and bedaquiline resistant | 4 (0.3%) | 3 (0.4%) | 1 (0.1%) |
| Extensively drug resistance | 2 (0.1%) | 2 (0.2%) | 0 |
| ***M. tuberculosis* lineage, n** |  |  |  |
| Lineage 1 | 69 (4.6%) | 40 (5.0%) | 29 (4.2%) |
| Lineage 2 | 1320 (88.5%) | 704 (87.8%) | 616 (89.4%) |
| Lineage 4 | 40 (2.7%) | 25 (3.1%) | 15 (2.2%) |
| Mixed or unknown | 62 (4.2%) | 33 (4.1%) | 29 (4.2%) |

We incorporated 2,922 previously published *M. tuberculosis* genomes from Ho Chi Minh City alongside 3,050 contemporary sequences generated in this and a parallel study to increase phylogenetic resolution and provide the ancestral context needed to detect the emergence of resistance in our phylogenetic analysis. After excluding 254 mixed-lineage sequences, the combined dataset comprised 5,718 genomes, half of which were rifampicin-susceptible **(Table S2, Figure S2)**. Model comparisons identified the TVM nucleotide substitution model as the best fit for reconstructing the phylogeny.

Among the data this study generated (64% sampling coverage), the N approach estimated that 19.5% (17.8 to 21.6%) of RR-TB was attributable to acquired resistance compared to 29.4% (27.7 to 31.6%) under N-1 approach. Among people with previous TB (under the N approach), 36% (34-39%) of RR-TB was estimated to arise through *de novo* acquisition and 64% (61-66%) was attributed to transmission. Extrapolating to complete sampling coverage using SIMEX, we estimated a plausible range for *de novo* acquired rifampicin resistance of 13 to 28%. In the sensitivity analysis, temporal depth of RR-TB sampling had minimal independent effect on classification when sample size was held constant, suggesting that starting sampling earlier than 2020 would not have substantially changed our estimates **(Supplementary Results)**.

Across the three major circulating lineages in Viet Nam (Lineages 1, 2, and 4), we identified 220 rifampicin-resistance emergence events, of which the *rpoB S450L* mutation was consistently the most frequent across lineages [122 (55%)]. The earliest inferred emergence occurred in Lineage 2 and was dated to 1980 (95% credible interval 1970–1989). Viet Nam’s NTP introduced rifampicin nationwide from 1986.^9^ For Lineages 1 and 4, the estimates were later, although the credible intervals were similarly wide. Subsequent emergence of resistance events were estimated to have continued to the present day **(Figure 3).**

**Figure 3:**
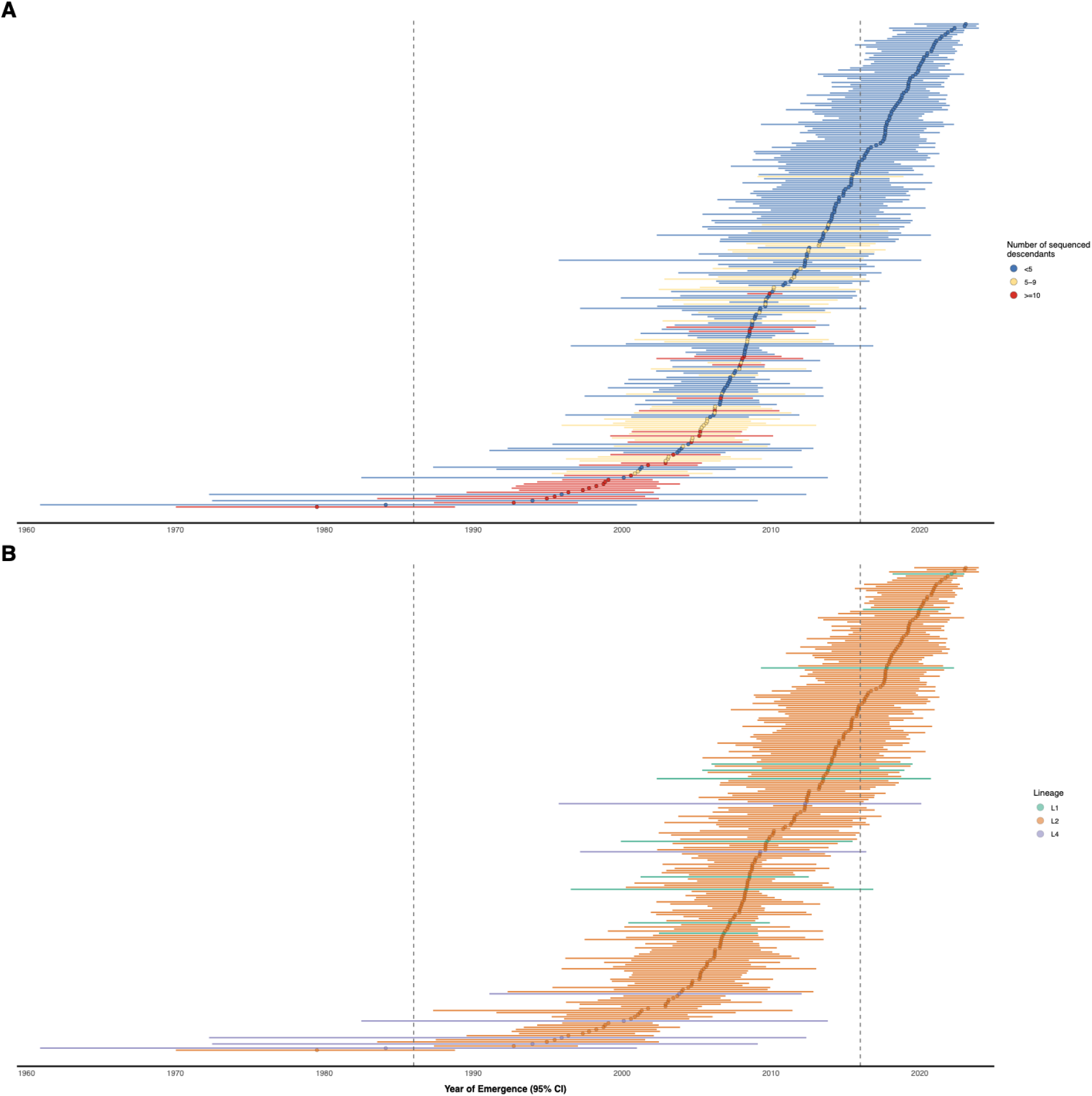
**Estimated dates of rifampicin-resistance emergence events for transmitted *M. tuberculosis* strains in Ho Chi Minh City** (A) Emergence events coloured by number of sequenced descendants. (B) Emergence events coloured by lineage. Dashed line at 1986 denotes the formation of the Viet Nam National Tuberculosis Programme and the nationwide rollout of rifampicin. Dashed line at 2016 denotes nationwide roll of Xpert MTB/RIF assay

**Figure 4:**
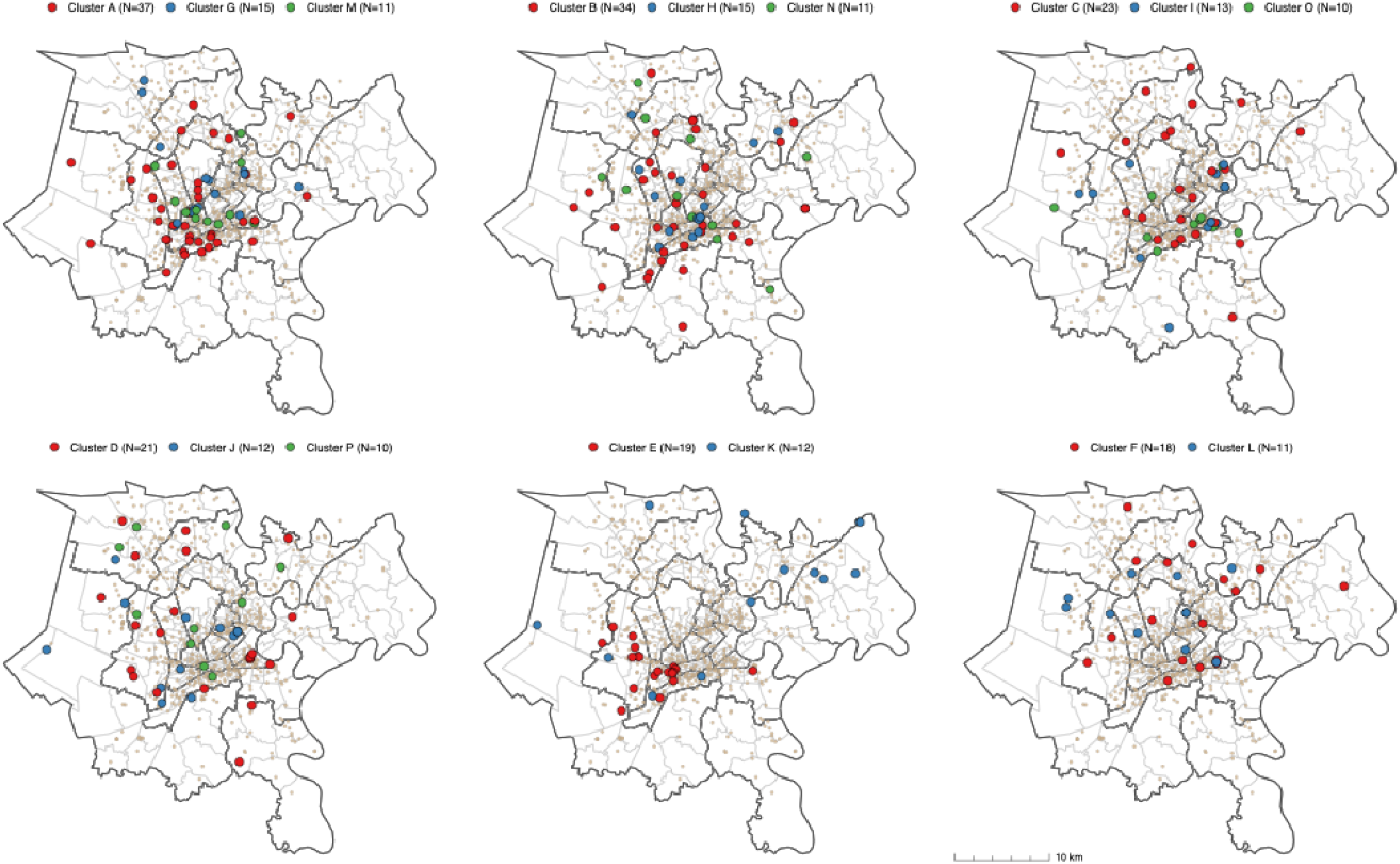
Geographic distribution of rifampicin-resistant tuberculosis transmission clades including ≥10 cases in Ho Chi Minh City. Tan coloured dots represent cases that are not members of any identified transmission network. Transmission networks are grouped across multiple copies of the Ho Chi Minh City map to avoid overcrowding any single map. Maps exclude Cần Giờ and Củ Chi districts as no cases from large transmission networks located to these districts.

When examining predictors of transmitted versus acquired rifampicin resistance (among participants with previous TB), we found no evidence that birthplace (Ho Chi Minh City versus elsewhere) influenced the inferred pathway to resistance (OR 0.92 [95% CI 0.69-1.23]. In contrast, infection with *M. tuberculosis* Lineage 2 was associated with lower odds of acquired (relative to transmitted) resistance (OR 0.23 [0.14–0.36]), whereas diabetes was associated with higher odds of acquired resistance (OR 1.58 [1.28–2.10]). No other host or pathogen covariates showed statistically significant associations **(Table S3)**.

We analysed transmission networks to estimate where in the city transmission was occurring. After excluding historic cases for whom residential addresses were unavailable, we identified 213 monophyletic RR-TB transmission networks, ranging in size from 2 to 37 cases. Large transmission networks (>10 members) exhibited distinct spatial distributions across the city’s urban core but were dispersed across multiple districts rather than concentrated within single geographic hotspots **(Figure 4)**. Geographic dispersion of transmission networks, measured as median pairwise commute time between cases, increased with time since network emergence (Spearman rho = −0.25, p = 0.002).

We identified 18 shared households comprising 38 participants (2.6% among 1,486 cases with valid addresses). 17 households contained two cases and one household contained four. In only 8/18 (44.4%) households did all cases belong to the same transmission network.

RR-TB transmission in Ho Chi Minh City was only weakly structured by host demographics or geography, with most genetic relatedness reflecting broad, city-wide transmission rather than assortative mixing within specific demographic or spatial subgroups. treeSeg detected no clades with distinct trait distributions for any binary demographic variable (sex, HIV status, diabetes, birthplace and drug use history). For age, treeSeg identified a single small clade of 13 isolates (0.9% of the sample) with a lower mean age (21 versus 46 years), likely reflecting a localised transmission chain rather than systematic age-dependent structuring. There was no correlation between genetic and geographic distance (Mantel r = 0.001, p = 0.94), and pairwise commute times were similar for within-network and between-network pairs (median 14.0 versus 14.6 minutes). While individual transmission networks (≥10 members) differed in their demographic composition for several characteristics **(Figure S3)**, this did not translate into assortative transmission at the population level where demographic and geographic similarity together explained little of the variance in genetic distance (MRM R² = 0.62%).

## Discussion

In this large, population-based genomic epidemiology study of RR-TB in Ho Chi Minh City, we show that transmission of already-resistant strains accounts for most of the current RR-TB burden, despite the fact that most patients with RR-TB have had a previous episode of TB. Nevertheless, a substantial and concerning proportion of RR-TB arises through *de novo* acquired resistance, despite long-standing programmatic efforts to prevent this.

We further reconstructed the evolutionary history of RR-TB in Ho Chi Minh City and found that rifampicin resistance likely emerged not long after the introduction of the drug in the mid-1980s. We demonstrate that at least some of those early resistance events have resulted in networks that have persisted ever since, and that are now widespread across the city. Whilst most contemporary RR-TB in Ho Chi Minh City is the product of transmission, we find that the emergence of rifampicin-resistance has continued since the 1980s, despite the maturation of the NTP and the nationwide rollout of Xpert MTB/RIF by 2016.^29^

The predominance of transmitted rifampicin resistance we observe aligns with findings from other high-burden settings. In a South African study, 70% of RR-TB isolates clustered within 5 SNPs and were attributed to transmission, while an analysis of more than 5,000 strains from China estimated that 61% of RR-TB arose through transmission using similar methods.^30,31^ Estimates from SNP-based clustering may, however, be sensitive to sampling density and assumptions regarding the independence of resistance events. Incomplete sampling is likely to underestimate transmission while failure to account for homoplasy can inflate transmission estimates, particularly for rifampicin where a small number of resistance mutations occur frequently.^22,32^ Several smaller studies have therefore built upon SNP-based approaches by applying phylogenetic methods, such as an analysis of 576 strains from Tibet which reported that 72% of MDR-TB was transmitted.^33^ Our framework integrates additional sources of information including prior TB history, N and N −1 classification approaches and correction for sampling coverage which may improve the distinction of transmitted from acquired rifampicin resistance, particularly among individuals without previous TB. We explicitly characterise uncertainty arising from sampling variability, phylogenetic inference, and classification boundaries, yielding estimates that more fully reflect the inherent limitations of inferring resistance origins from these data.

The predominance of transmitted resistance, even among people with previous TB, and the diffuse spatial structure of large RR-TB transmission networks may have important implications for TB control in urban Viet Nam. We previously showed, using routine case notifications, that Ho Chi Minh City’s central districts were RR-TB incidence hotspots.^10^ The present genomic analysis provides finer resolution.

Although some large transmission networks are anchored in high-incidence districts, most are geographically dispersed, spanning multiple districts rather than forming tightly localised hotspots. This pattern may suggest that RR-TB transmission in Ho Chi Minh City is not primarily driven by household spread or small neighbourhood clusters, but instead reflects broader city-wide transmission. Indeed, despite most residents living in multi-person households, we identified only 18 affected households, less than half of which showed evidence of infection with the same strains. Nevertheless, inferences about transmission locations based solely on individuals with symptomatic disease are inherently uncertain given the complex natural history of TB, in which transmission may occur long before clinical presentation and many newly infected individuals may never progress to symptomatic disease.^34^ Ultimately, our findings support the prioritisation of broad, population-level interventions aimed at interrupting transmission to achieve the greatest reductions in RR-TB burden. While community-wide screening is inherently resource-intensive, it has been shown to be highly effective in other Vietnamese settings and is likely to be cost-effective over the long term.^3,35^

Between 13 and 28% of RR-TB in our study was attributable to acquired resistance, a substantial figure given the long-standing DOTS implementation and high first-line treatment success rates in our setting.^36^ This finding suggests that factors beyond incomplete or inadequate treatment may be contributing to the emergence of rifampicin resistance. One plausible mechanism is unrecognised isoniazid resistance among people with rifampicin-susceptible TB. In these patients, standard first-line therapy provides only three effective drugs during the intensive phase and may functionally reduce to rifampicin monotherapy during the continuation phase. In a large systematic review, the treatment of isoniazid-resistant, rifampicin-susceptible TB with standard first-line regimens was associated with a 25-fold increased risk of acquiring multidrug resistance compared to isoniazid-susceptible disease.^37^ Isoniazid mono-resistance may affect up to 20% of new TB cases in Viet Nam^38^, more than double the global average^39^, potentially creating a substantial reservoir for the amplification of rifampicin resistance. Host factors may also contribute, such as NAT2 fast-acetylator genotypes that reduce isoniazid exposure. In a small Vietnamese cohort, one-quarter of individuals carried fast-acetylator genotypes.^40^ Finally, we observed higher odds of acquired resistance amongst people with diabetes, consistent with prior reports, although underlying mechanisms remain uncertain.^41^

Our study has several limitations. Although we achieved substantially higher sampling coverage than most population-based TB studies from high-burden settings, interruptions during the COVID-19 pandemic and culture or sequencing failures meant that one-third of RR-TB cases were unsampled. While we attempted to correct for this using SIMEX, the correction assumed unsampled cases were randomly distributed but systematic differences in which patients underwent culture and sequencing could have introduced residual bias. Our classification of transmitted versus acquired resistance integrated phylogenetic evidence and clinical history, but there may still have been residual misclassification as treatment histories may have been incomplete, and infection outside Ho Chi Minh City could not be fully excluded, although these cases would still be correctly classified as transmitted if not previously treated for TB. Enrolment was restricted to cases detected through programmatic Xpert testing and initiating treatment at Phạm Ngọc Thạch Hospital excluding asymptomatic individuals and those who died before diagnosis. Finally, although our findings are context-specific and may not generalise, the analytic framework we describe should be applicable to similar investigations elsewhere.

RR-TB in Ho Chi Minh City is driven predominantly by ongoing transmission but a substantial contribution from *de novo* acquired resistance signals persistent vulnerabilities in current diagnostic and treatment pathways. Intensified efforts to interrupt transmission through earlier case detection and rapid treatment initiation will need to be coupled with further investigation into the precise drivers of *de novo* acquired drug-resistance.

## Data sharing

All newly generated sequences will be made available in the European Nucleotide Archive under study accession PRJEB107150 on final publication of the study.

Accession numbers for previously published sequences are provided in the supplementary files.

## Data Availability

All newly generated sequences will be made available in the European Nucleotide Archive under study accession PRJEB107150 on final publication of the study. Accession numbers for previously published sequences are provided in the supplementary files.

## Acknowledgements

RS is supported by the Rhodes Trust. This research was funded in whole, or in part, by the Wellcome Trust (214560/Z/18/Z). TMW is a Wellcome Trust Clinical Career Development Fellow (214560/Z/18/Z).

## Declaration of generative AI and AI-assisted technologies in the manuscript preparation process

During the preparation of this work the author(s) used Claude AI in order to improve language and readability. After using this tool, the author(s) reviewed and edited the content as needed and take(s) full responsibility for the content of the published article.

