## Supplementary Methods for "Transmission of rifampicin-resistant tuberculosis in Ho Chi Minh City, Viet Nam: a prospective genomic epidemiology study"

**Supplementary appendix**

### Supplementary methods

#### DNA extraction, library preparation and whole genome sequencing

Mature colonies were harvested and suspended in 400 µL of 1× Tris-EDTA (TE) buffer (Thermo Fisher Scientific, Waltham, MA, USA). The suspensions were sonicated at 35 kHz for 15 minutes and subsequently heated at 80 °C for 20 minutes to inactivate cells. After cooling to room temperature, 50 µL of lysozyme (10 mg/mL; Sigma-Aldrich, St. Louis, MO, USA) was added, and samples were incubated overnight at 37 °C. Genomic DNA was then extracted using the cetyl trimethylammonium bromide (CTAB) method.^1^ Extracted DNA was prepared for sequencing using a modified Nextera XT library preparation protocol,^2^ generating 2 × 150 bp paired-end reads. Pooled libraries were sequenced on Illumina NextSeq 1000/2000 platforms, aiming for a mean depth of at least 50X.

#### Additional *M. tuberculosis* sequences

Previously published genomes

Previously published WGS datasets from Ho Chi Minh City were incorporated to maximise phylogenetic resolution. The foundational dataset comprised 1,635 M.tuberculosis isolates collected from HIV-negative, smear-positive adults initiating treatment at district TB units between 2008 and 2011, as described by Holt et al.^3^ This collection was subsequently expanded by Silcocks et al. who analysed a total of 2,542 genomes from Ho Chi Minh City TB patients recruited between 2001 and 2013, incorporating the majority of the Holt et al. pulmonary isolates and adding newly sequenced pulmonary and TB meningitis isolates.^4^ We additionally included Ho Chi Minh City isolates contributed through the CRyPTIC consortium. These comprised pulmonary and extrapulmonary clinical isolates collected retrospectively from biobanked specimens between 2009 and 2019.

Genomes generated in parallel study

Two case–control studies conducted in parallel with the prospective RR-TB cohort study, between March 2020 and April 2024, contributed additional *M. tuberculosis* genomes to the combined Ho Chi Minh City dataset. Both studies analysed participants with a first episode of Xpert-confirmed rifampicin-susceptible pulmonary TB (PTB) and followed them longitudinally to identify microbiologically confirmed recurrence. For both studies, isolates from the baseline episode and, where applicable, the recurrence episode were included to maximise phylogenetic resolution and to improve detection of resistance-emergence events. In the **first study**, controls were individuals with rifampicin-susceptible PTB who completed treatment without recurrence during the study period. Cases were individuals who experienced **rifampicin-susceptible TB recurrence**. In the **second study**, cases were individuals with an initial rifampicin-susceptible PTB episode who subsequently developed **rifampicin-resistant TB recurrence**. **Here recurrence isolates** contributed directly to the present RR-TB cohort, which aimed to include all Xpert-confirmed RR-TB cases city-wide.

#### Phylogenetic analyses

Fixed-length FASTA files for individual isolates generated by the GPAS^5^ pipeline were combined into multi-sequence SNP alignments with runListCompare v0.3.8 (<https://github.com/davideyre/runListCompare>). Alignment positions not called (labelled as N) in more than 10% of sequences were excluded, and repetitive and low-complexity regions of the genome were masked to reduce the influence of poorly supported or unreliable sites on phylogenetic inference. SNP alignments were used to generate maximum-likelihood phylogenetic trees using IQ-TREEv3.0.1.^6^ ModelFinder was used to identify the best substitution model.^7^ To correct for ascertainment bias, the number of invariant A, C, G, and T sites in the full alignment was specified using the
**-fconst** option. For the analysis of acquired versus transmitted resistance, phylogenies were constructed employing the -fast option in IQ-TREE to allow efficiency across the large number of bootstrap replicates. Phylogenetic trees were visualised with Interactive Tree of Life (iTOL) v7.2.2.^8^

Simulation-extrapolation (SIMEX) correction for incomplete sampling
Incomplete sampling may bias phylogenetic classification of transmitted versus acquired resistance, leading to systematic underestimation of transmission. When RR-TB cases are missing from the sample, true transmission clusters may be fragmented, causing mutations that occurred on internal branches (indicating transmission) to instead appear as independent tip-level homoplasies (suggesting acquisition). To correct for this bias, we applied a simulation-extrapolation (SIMEX) framework.^9^

We generated datasets at ten sampling fractions representing 5% to 59% of the estimated true RR-TB population (corresponding to keeping 8% to 92% of our observed sample). At each fraction, we created five independent replicates by randomly subsampling (without replacement) RR-TB isolates from the present study while retaining all context isolates to preserve phylogenetic structure. For each subsampled dataset, we inferred a maximum-likelihood phylogeny, mapped rpoB mutations using SNPPar^10^, and classified resistance under both the N and N-1 classification approaches. Combined with our observed data at 64% sampling, this yielded 51 phylogenies spanning the sampling fraction range.

To extrapolate to complete sampling, we fitted generalized additive models (GAMs) with binomial errors and logit link to the relationship between sampling fraction (λ) and the proportion of RR-TB due to transmission. The model took the form: logit(transmitted) = f(λ) where f(λ) is a thin-plate regression spline. We fitted separate models for the N and N-1 approaches. Smoothness selection was performed using restricted maximum likelihood (REML), which penalises overfitting; model adequacy was therefore assessed using the k-index diagnostic to ensure sufficient basis dimension (i.e., no underfitting). We then extrapolated each fitted curve to λ = 1.0 (complete sampling) to obtain SIMEX-corrected estimates. Confidence intervals were derived from the standard errors of the GAM predictions on the link scale, back-transformed to the probability scale. Convergence of estimates across replicates confirmed that five replicates per sampling fraction was sufficient for stable estimation.

Sensitivity analysis for temporal depth of sampling

To assess whether estimates would differ had RR-TB sampling begun earlier than 2020, we performed a sensitivity analysis that isolated the effect of temporal depth from sample size. For each year range (2024 only, 2023–2024, 2022–2024, 2021–2024, and 2020–2024), we randomly subsampled RR-TB isolates to a fixed sample size equal to the minimum available in any single year, while retaining all context isolates. We generated five replicates per year range, rebuilt phylogenies, and classified resistance under both approaches. We then fitted GAMs with number of years as the predictor to assess whether temporal depth independently influenced classification after controlling for sample size.

#### Temporal emergence of rifampicin resistance

Clock rate estimation

Running BactDating^11^ with clock rate estimation is computationally intensive on large phylogenies. We therefore developed a subsampling approach to obtain context-specific clock rate estimates while maintaining computational tractability. For each lineage, we generated 20 representative subsamples of 200 isolates using stratified temporal sampling. Isolates were allocated across 10 time bins based on collection date quantiles, with uniform random sampling within each bin, to maintain the temporal distribution of the full dataset without biasing toward outliers that could inflate rate estimates.

For each subsample, maximum-likelihood phylogenies were reconstructed using IQ-TREE with the TVM substitution model, 1,000 ultrafast bootstrap replicates, and correction for invariant sites. In addition to repetitive and low-complexity regions, drug-resistance associated genes were also masked. Trees were rooted by optimising root-to-tip regression using BactDating's initRoot function. BactDating was then run under an additive uncorrelated relaxed-clock model^12^ with clock rate estimation enabled, using previously published lineage-specific rates as informative priors: 1.07 substitutions/genome/year for Lineage 1, 0.538 for Lineage 2 and 0.237 for Lineage 4.^13^ MCMC sampling was run for 5 × 10^7^ iterations.

Posterior distributions of clock rates were pooled across replicates with adequate convergence (effective sample size ≥ 100 for the mutation rate parameter). The pooled median rate was used as a fixed clock rate for dating the full phylogeny.

Clock rate estimates for Lineages 1 and 2 were 1.866 (0.777–3.830) and 0.876 (0.446–2.120) substitutions/genome/year, respectively. For Lineage 4, none of the 20 subsampled replicates achieved adequate convergence (ESS < 100 for all replicates), suggesting insufficient temporal signal to reliably estimate a context-specific rate. We therefore applied the published Lineage 4 rate of 0.237 substitutions/genome/year as a fixed clock rate.

Full phylogeny dating
For each lineage, a maximum-likelihood phylogeny of the full dataset was inferred using IQ-TREE, this time unmasking resistance associated genes from the alignment, and rooted using temporal signal optimisation. Branch lengths were scaled to substitutions per genome by multiplying by the M. tuberculosis H37Rv reference genome length (4,411,532 bp).

BactDating was run under an additive uncorrelated relaxed-clock model with fixed lineage-specific clock rates: the estimated rates for Lineages 1 and 2, and the published rate for Lineage 4. MCMC sampling was run for 1 × 10^6^ iterations. Convergence was confirmed by inspection of trace plots and effective sample sizes > 200 for alpha and sigma parameters.

Temporal signal assessment
Temporal signal was assessed using ten date-randomisation tests per Lineage, in which sampling dates were permuted among isolates. Model fit was compared using the Deviance Information Criterion (DIC). Lower DIC values for the true-dated analyses relative to randomised replicates were interpreted as evidence of sufficient temporal signal.^11^

Resistance emergence dating
Homoplastic rpoB mutations conferring rifampicin resistance were identified from the rooted maximum-likelihood phylogenies using SNPPar.^10^ The timing of rifampicin-resistance emergence was inferred from posterior node ages corresponding to the most recent common ancestors of clades carrying these homoplastic resistance mutations.

Geographic and demographic structure of RR-TB transmission networks
To test for assortative transmission of host characteristics, we used treeSeg^14^ to test whether the distribution of each trait depended on phylogenetic position. treeSeg uses a multiscale change-point framework that simultaneously tests for clades with distinct trait distributions at all levels of the tree hierarchy while controlling the overall false positive rate at a prespecified level α. For a given α, the method identifies ‘active nodes’ (internal nodes where the trait distribution changes) and provides confidence sets for their locations. We applied treeSeg at α = 0.05 to each binary trait (sex, HIV status, diabetes, birthplace and, drug use history) and to age as a continuous variable under a Gaussian model. The tree was midpoint-rooted and polytomies were resolved randomly prior to analysis.

To assess whether RR-TB transmission was spatially structured or panmictic, we evaluated the relationship between genetic relatedness and geographic proximity using two approaches. Geographic proximity was measured as commute time (minutes) calculated using the Open-Source Routing Machine (OSRM), based on OpenStreetMap road network data.^15^ Pairwise commute times were extracted for all sample pairs with available coordinate data and symmetrised. First, we performed a Mantel test correlating pairwise phylogenetic distances with pairwise commute times. Pearson's correlation coefficient was used, and statistical significance was assessed using 1,000 permutations. Second, we compared commute times between isolate pairs within the same transmission network versus pairs from different networks. Commute times were summarised using medians and interquartile ranges, and within- versus between-network comparisons were performed using the Wilcoxon rank-sum test.

To quantify the joint contribution of demographic similarity and geography to genetic relatedness while accounting for correlations among predictors, we performed Multiple Regression on Distance Matrices (MRM). Pairwise phylogenetic distance matrices were constructed for each predictor. For binary variables, distances were defined as 0 for concordant pairs and 1 for discordant pairs. For age, Euclidean distances were calculated on standardised values. Commute time (minutes) derived from OSRM was used as the geographic distance matrix. We fitted an MRM model including demographic predictors and commute time, restricted to isolates with complete routing data. Models were fitted using permutation-based inference (1,000 permutations). Model fit was summarised using the coefficient of determination (R²), which represents the proportion of variance in phylogenetic distance explained by the predictors.

### Supplementary results

#### SIMEX correction for incomplete sampling

Both GAM models demonstrated excellent fit to the simulated data **(Figure S4)**. The N approach model explained 82.1% of deviance with effective degrees of freedom (edf) of 1.6, indicating a near-linear relationship between sampling fraction and transmission estimates. The N-1 method model explained 86.6% of deviance with edf of 2.5, indicating slightly more curvature in the bias-correction relationship. The k-index diagnostics (1.08 and 1.05 respectively, both p > 0.5) confirmed adequate basis dimension for both models, indicating no underfitting. Convergence assessment showed that estimates stabilised within 3–5 replicates at each sampling fraction.

Under the N approach, the SIMEX-corrected estimate of acquired resistance was 13.3% (10.9–16.2%), a correction of 6.2 percentage points. Under the N-1 approach, the corrected estimate was 28.1% (22.2–35.0%), a correction of 1.3 percentage points.

Sensitivity analysis for temporal depth of sampling

When sample size was held constant (N=123) while varying temporal depth from 1 to 5 years, the N approach showed no relationship between temporal depth and the estimated proportion of transmitted resistance (5.9% deviance explained; estimates ranged from 72.4% to 77.9% with no trend). The N-1 approach showed a modest downward trend (30.5% deviance explained), with transmission estimates decreasing from 59.7% at 1 year to 51.5% at 5 years, the opposite direction to what would be expected if additional historical context improved identification of transmission links **(Table S5)**. The absence of a positive temporal effect after controlling for sample size suggests that the bias identified by SIMEX is driven primarily by incomplete cross-sectional sampling rather than limited historical context, and that starting RR-TB sampling earlier than 2020 would not have substantially changed our estimates.

### Supplementary tables

#### Table S1: Demographic characteristics of enrolled participants with Xpert-confirmed rifampicin-resistant tuberculosis, stratified by whole-genome sequencing status

|  | No isolate sequenced (N=334) | Isolate sequenced (N=1699) | Overall (N=2033) |
| --- | --- | --- | --- |
| Age, mean (SD) | 47.6 (14.9) | 46.0 (14.1) | 46.3 (14.3) |
| Male sex, n | 250 (74.9%) | 1226 (72.2%) | 1476 (72.6%) |
| Body mass index, mean (SD) | 19.8 (3.45) | 19.5 (3.22) | 19.5 (3.26) |
| Birthplace Ho Chi Minh City, n | 224 (67.1%) | 1125 (66.2%) | 1349 (66.4%) |
| Previous TB episode, n | 188 (56.3%) | 922 (54.3%) | 1110 (54.6%) |
| Previous RR-TB, n | 16 (4.8%) | 77 (4.5%) | 93 (4.6%) |
| Smear positive TB | 154 (46.1%) | 1256 (73.9%) | 1410 (69.4%) |
| HIV positive | 24 (7.2%) | 104 (6.1%) | 128 (6.3%) |
| Diabetes | 101 (30.2%) | 500 (29.4%) | 601 (29.6%) |

#### Table S2: Resistance classification and lineage distribution of contemporary and previously published genomes included in analysis of acquired versus transmitted resistance

|  | **Contemporary genomes (N=2870)** | **Previously published genomes (N=2848)** | **Overall (N=5718)** |
| --- | --- | --- | --- |
| **Resistance classification** |  |  |  |
| Rifampicin and isoniazid suspectable | 977 (34.0%) | 1852 (65.0%) | 2829 (49.5%) |
| Isoniazid mono-resistant | 292 (10.2%) | 676 (23.7%) | 968 (16.9%) |
| Rifampicin mono-resistant | 162 (5.6%) | 21 (0.7%) | 183 (3.2%) |
| Multidrug resistant | 1198 (41.7%) | 282 (9.9%) | 1480 (25.9%) |
| Pre-extensively drug resistant | 234 (8.2%) | 17 (0.6%) | 251 (4.4%) |
| Multidrug and bedaquiline resistant | 4 (0.1%) | 0 (0%) | 4 (0.1%) |
| Extensively drug resistant | 3 (0.1%) | 0 (0%) | 3 (0.1%) |
| ***M. tuberculosis* lineage, n** |  |  |  |
| Lineage 1 | 319 (11.1%) | 631 (22.2%) | 950 (16.6%) |
| Lineage 2 | 2430 (84.7%) | 1922 (67.5%) | 4352 (76.1%) |
| Lineage 3 | 2 (0.1%) | 3 (0.1%) | 5 (0.1%) |
| Lineage 4 | 119 (4.1%) | 292 (10.3%) | 411 (7.2%) |

#### Table S3: Univariate associations with acquired versus transmitted rifampicin resistance among participants with previous tuberculosis

| **Variable** | **Odds Ratio (95% CI)** |
| --- | --- |
| Lineage 2 Other | 0.23 (0.14-0.36) Reference |
| Age (per year) | 1.01 (1.00-1.02) |
| Male Female | 0.92 (0.66-1.26) Reference |
| Body mass index (per kg/m^2^) | 1.04 (0.99-1.08) |
| Number of previous TB episodes (per episode) | 1.25 (0.94-1.67) |
| HIV positive HIV negative | 0.95 (0.58-1.50) Reference |
| Diabetes No Diabetes | 1.58 (1.18-2.10) Reference |
| Previous smoker Current smoker Never smoker | 1.12 (0.82-1.53) 1.21 (0.85-1.71) Reference |
| Alcohol <14 units per week Alcohol ≥14 units per week No alcohol | 1.05 (0.77-1.44) 1.00 (0.72-1.49) Reference |
| Previous intravenous drug use No intravenous drug use | 0.60 (0.30-1.14) Reference |
| Born in Ho Chi Minh City  Born outside Ho Chi Minh City | 0.92 (0.69-1.23)  Reference |

### Supplementary figures

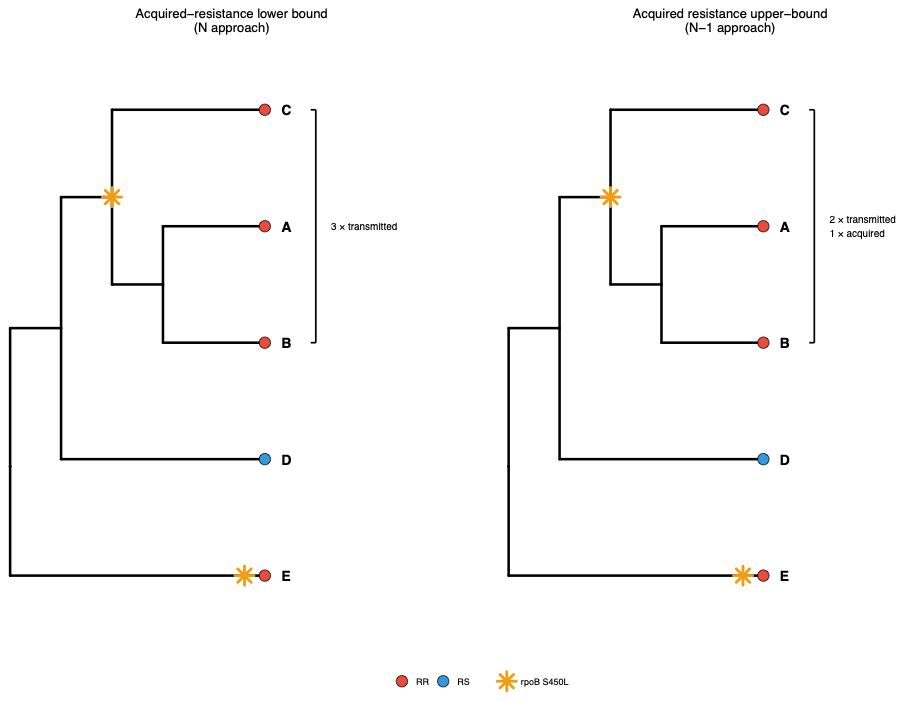

#### Figure S1: Schematic illustration of N and N-1 classification approaches for acquired rifampicin resistance

Schematic phylogeny showing five sampled *Mycobacterium tuberculosis* isolates (A–E) with rifampicin-resistant (RR) and rifampicin-susceptible (RS) phenotypes indicated. The emergence of rifampicin resistance is indicated by a homoplastic *rpoB* S450L mutation, represented by the yellow star. When such a resistance-conferring mutation maps to an internal node of the phylogeny, its origin cannot be uniquely attributed to transmission or within-host acquisition. Under the N approach, all descendants of an internal resistance-associated node (A, B & C) are classified as having transmitted resistance, resulting in three transmitted RR cases in this example. Under the N-1 approach, the presence of at least one previously treated individual among the descendants permits one event to be classified as acquired resistance and the remaining as transmitted. These approaches therefore define plausible lower and upper bounds on the proportion of rifampicin-resistant TB attributable to acquired resistance in the presence of phylogenetic ambiguity. Isolate E represents a tip-level homoplasy classified as acquired resistance under both approaches.

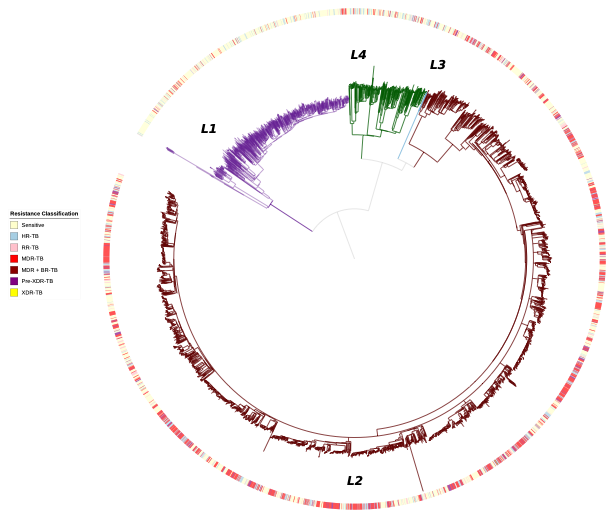

#### Figure S2: Consensus maximum-likelihood phylogenetic tree for 5,718 M.tuberculosis isolates from Ho Chi Minh City, Viet Nam

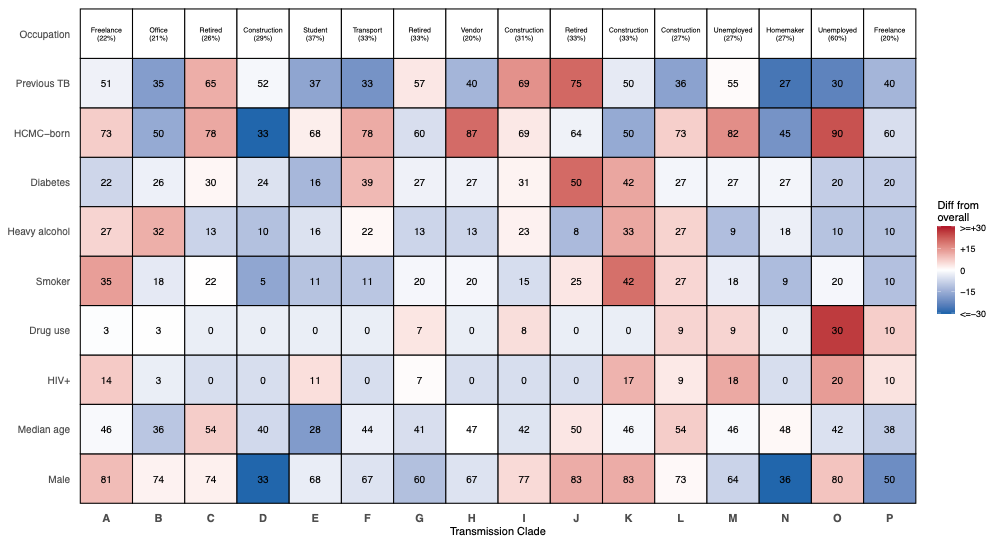

#### Figure S3: Occupational and demographic profiles for large RR-TB transmission clades.

Heatmap illustrating compositional heterogeneity across RR-TB transmission networks with ≥10 members (A–P). For each network, the top row indicates the most common occupation and its proportion among members. Cell values represent percentages for categorical variables and median values for age (y-axis labels). Cell colours denote the magnitude and direction of deviation (percentage points) from the overall RR-TB population, with red indicating higher and blue indicating lower values relative to the overall distribution.

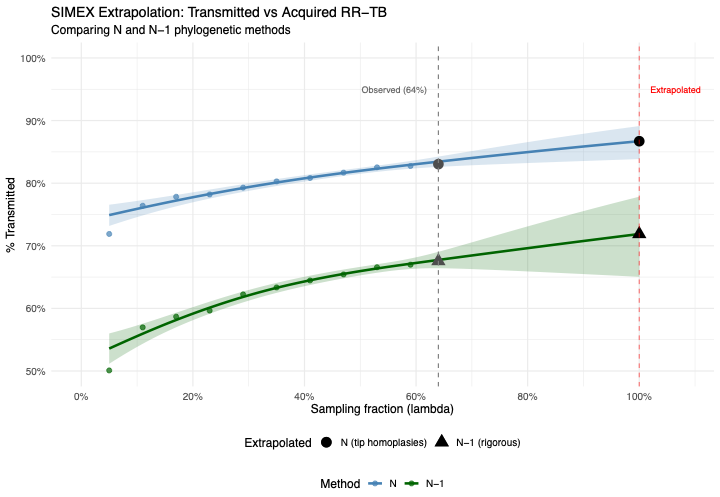

#### Figure S4. Simulation–extrapolation (SIMEX) correction for incomplete sampling in estimates of transmitted versus acquired rifampicin-resistant tuberculosis

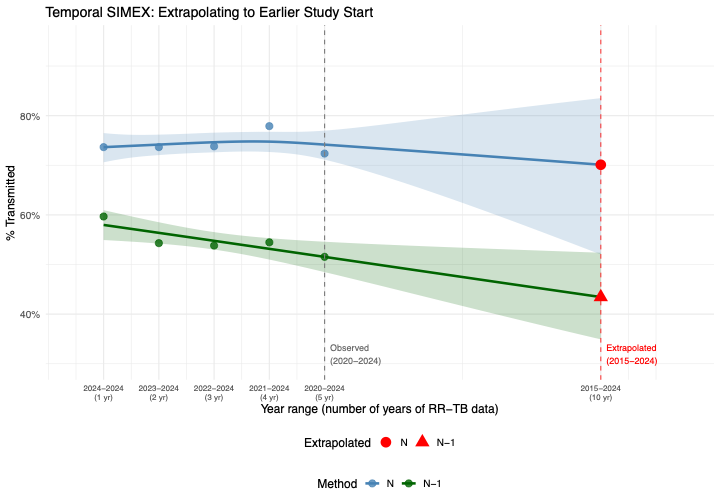

#### Figure S5. Simulation–extrapolation (SIMEX) correction for temporal depth in estimates of transmitted versus acquired rifampicin-resistant tuberculosis
